# Cross-reactive CD4^+^ T cells enhance SARS-CoV-2 immune responses upon infection and vaccination

**DOI:** 10.1101/2021.04.01.21252379

**Authors:** Lucie Loyal, Julian Braun, Larissa Henze, Beate Kruse, Manuela Dingeldey, Ulf Reimer, Florian Kern, Tatjana Schwarz, Maike Mangold, Clara Unger, Friederike Dörfler, Shirin Kadler, Jennifer Rosowski, Kübrah Gürcan, Zehra Uyar-Aydin, Marco Frentsch, Florian Kurth, Karsten Schnatbaum, Maren Eckey, Stefan Hippenstiel, Andreas Hocke, Marcel A. Müller, Birgit Sawitzki, Stefan Miltenyi, Friedemann Paul, Marcus A. Mall, Holger Wenschuh, Sebastian Voigt, Christian Drosten, Roland Lauster, Nils Lachman, Leif-Erik Sander, Victor M. Corman, Jobst Röhmel, Lil Meyer-Arndt, Andreas Thiel, Claudia Giesecke-Thiel, the Charité Corona Cross Study Group

**Author notes:** These authors contributed equally to this work (Julian Braun, Larissa Henze, Beate Kruse, Manuela Dingeldey, Ulf Reimer, Florian Kern, Tatjana Schwarz). These authors jointly supervised this work (Roland Lauster, Nils Lachman, Leif-Erik Sander, Victor M. Corman, Jobst Röhmel, Lil Meyer-Arndt, Andreas Thiel, Claudia Giesecke-Thiel).

## Abstract

While evidence for pre-existing SARS-CoV-2-cross-reactive CD4^+^ T cells in unexposed individuals is increasing, their functional significance remains unclear. Here, we comprehensively determined SARS-CoV-2-cross-reactivity and human coronavirus-reactivity in unexposed individuals. SARS-CoV-2-cross-reactive CD4^+^ T cells were ubiquitous, but their presence decreased with age. Within the spike glycoprotein fusion domain, we identified a universal immunodominant coronavirus-specific peptide epitope (*iCope*). Pre-existing spike- and *iCope*-reactive memory T cells were efficiently recruited into mild SARS-CoV-2 infections and their abundance correlated with higher IgG titers. Importantly, the cells were also reactivated after primary BNT162b2 COVID-19 mRNA vaccination in which their kinetics resembled that of secondary immune responses. Our results highlight the functional importance of pre-existing spike-cross-reactive T cells in SARS-CoV-2 infection and vaccination. Abundant spike-specific cross-immunity may be responsible for the unexpectedly high efficacy of current vaccines even with single doses and the high rate of asymptomatic/mild infection courses.

---

The newly emerged severe acute respiratory syndrome coronavirus 2 (SARS-CoV-2) caused a global pandemic challenging economy and health systems. In the majority of infected individuals, coronavirus disease 2019 (COVID-19) manifests in asymptomatic or mild phenotypes. However, severe and fatal disease occurs in about 5% of those infected and are primarily associated with age and comorbidities such as cardiovascular, pulmonary and kidney diseases and diabetes (*1*). Initially, it was assumed that SARS-CoV-2 encountered an immunologically unprotected population, however, SARS-CoV-2 displays considerable homologies with endemic, seasonal common cold endemic coronaviruses (collectively referred to as ‘HCoV’) (*2, 3*). There is now firm evidence for the cellular and humoral cross-reactivity to SARS-CoV-2 (*3*-*14*), although the role of cross-reactive immunity in SARS-CoV-2 infection is controversial, and potential negative effects have been discussed (*2, 8, 15, 16*). However, it has been reported that recent HCoV infection is associated with less severe COVID-19, suggesting a protective role (*17*). Thus, a better understanding of the extent and impact of cross-immunity in SARS-CoV-2 infection and vaccination is critical, as cognate cross-immunity would influence a vaccination regimen which should then be adjusted to achieve global population immunity in the most expeditious manner.

Here we determined the functional role of pre-existing SARS-CoV-2 and HCoV-reactive CD4^+^ T cells at a previously unattained resolution. We demonstrate broad T cell cross-reactivity in unexposed individuals with spike glycoprotein (spike) serving as one of the immunodominant targets. This cross-immunity decreased with age. Importantly, we identified an ***i***mmunodominant ***Co***ronavirus ***pe***ptide ***e***pitope (*iCope*) located within the fusion domain of spike recognized by CD4^+^ T cells in 20% of unexposed individuals, 50-60% of SARS-CoV-2 convalescents and 97% of BNT162b2 vaccinated individuals. *iCope* and spike-cross-reactive T cells were recruited into primary SARS-CoV-2 infection immune responses and also profoundly into BNT162b2 COVID-19 mRNA vaccination responses. Finally, cross-reactive immunity exhibited secondary response kinetics upon primary vaccination and abundance of pre-existing cross-reactive T cells correlated with high functional avidity already early in the immune response as well as with the early induction and stabilization of S1 IgG antibody titers. Our results demonstrate a role of cross-reactive CD4^+^ T cells in SARS-CoV-2 infection and vaccination and add new information on the discussion of implementing both single-dose vaccination of healthy adults and two-dose vaccination of the elderly.

## Frequent and broad SARS-CoV-2-cross-reactivity in unexposed healthy donors

To determine the extent of cellular cross-reactivity, we stimulated CD4^+^ T cells of 60 unexposed healthy donors and 59 COVID-19 convalescents as controls (Table 1) with the complete SARS-CoV-2 orfeome (Fig. 1A). The SARS-CoV-2 orfeome consists of 11 ORFs (NCBI), five of which (ORF1a/b, NCAP, spike, VEMP and VME) are also found in HCoVs 229E, OC43, NL63 and HKU1, with ORF1a/b coding for a total of 16 non-structural proteins (NSPs). Amino acid sequence alignment revealed discrete stretches of high homology in almost all SARS-CoV-2 proteins to the corresponding proteins in HCoVs. Parts of the ORF1a/b including NSP8, NSP10 and NSP12-16 displayed the highest frequencies of homology and thus potential cross-reactive epitopes to all HCoVs (Fig. S1a). COVID-19 convalescents displayed CD4^+^ T cells reacting strongly against spike N-terminal S-I (amino acid residues 1-643) and C-terminal S-II (amino acid residues 633-1273), NCAP, and VME peptide pools (Fig. 1A, B). However, there was no increased reactivity among convalescents compared to SARS-CoV-2-unexposed individuals with respect to NSP8-16, some of which showed the highest degree of sequence homology between HCoVs and SARS-CoV-2. Reactivity against the combination of S-I, S-II, NCAP, and VME peptide pools clearly distinguished unexposed individuals from COVID-19 convalescents irrespective of the disease course (Fig. 1C). In unexposed individuals, we detected low CD4^+^ T cell reactivity against virtually all SARS-CoV-2 antigens to various extent, including those exclusive to SARS-CoV-2 (Fig. S1b), however, there was no obvious correlation between homology and cross-reactivity across the proteins (Fig. 1D). Thus, we identified cognate cross-reactivity likely to result from previous exposure to similar proteins found in HCoVs with respect to, for example, the spike protein, but also some non-cognate cross-reactivity, i.e. cross-reactivity that cannot be explained by the previous exposure to similar proteins in HCoVs. Of all 30 orfeome peptide pools, spike S-I/-II was the only one that elicited T-cell reactivity in all COVID-19 convalescents and additionally responses in some of the unexposed individuals. In addition, antibodies to spike induced by SARS-CoV-2 infection can neutralize the virus (*18*), and all of the recently approved and highly effective SARS-CoV-2 vaccines includes the spike as antigen. Therefore, we next assessed pre-existing SARS-CoV-2 spike-cross-reactive T cells in unexposed individuals in more detail.

**Table 1:** Donor characteristics. p.s.o. = post symptom onset

| Cohort | Subcohort | Number donors | Age mean (Std.d.) | Age range | Gender (f/m) | ICU | Hospitalization | Mild/asymptomatic | Measurement day p.s.o. mean (Std.d.) | Measurement day p.s.o. range | Figure(s) |
| --- | --- | --- | --- | --- | --- | --- | --- | --- | --- | --- | --- |
| **Orfeome** |  |  |  |  |  |  |  |  |  |  | Fig. 1, S1 |
|  | COVID-19 convalescents | 59 | 50.6 (15.0) | 21-79 | 24/35 | 20 | 17 | 22 | 152 (57) | 17-262 |  |
|  | Unexposed | 60 | 45.7 (16.1) | 25-76 | 36/24 | - | - | - | - | - |  |
| **Basic CCC** |  |  |  |  |  |  |  |  |  |  | Fig. 2, 3, S3 |
|  | COVID-19 WHO 1-2 | 133 | 43.8 (13.6) | 18-78 | 85/48 | - | - | 133 | 128 (74) | 21-304 |  |
|  | COVID-19 WHO 3-4 | 19 | 53.1 (12.8) | 31-75 | 7/12 | - | 19 | - | 137 (41) | 50-214 |  |
|  | COVID-19 WHO 5-7 | 22 | 59.2 (13.3) | 27-79 | 6/16 | 22 | - | - | 143 (59) | 61-229 |  |
|  | Unexposed | 568 | 47.5 (11.8) | 18-98 | 439/129 | - | - | - | - | - |  |
| **Other pathogen pools** |  |  |  |  |  |  |  |  |  |  | Fig. 2 |
|  | Young (<30) | 20 | 23.5 (2.4) | 20-29 | 11/9 | - | - | - | - | - |  |
|  | Old (>65) | 19 | 70.4 (3.8) | 65-76 | 11/8 | - | - | - | - | - |  |
| **Follow-up** |  |  |  |  |  |  |  |  |  |  | Fig. 5 |
|  | COVID-19 convalescents | 17 | 39.2 (10.8) | 22-58 | 11/6 |  |  | 17 | see Table 2 | see Table 2 |  |
| **Vaccination** |  |  |  |  |  |  |  |  |  |  |  |
|  | Volunteers | 31 | 40.9 (11.5) | 20-67 | 12/17 | - | - | - | - | - | Fig. 6, S5 |

**Fig. 1:**
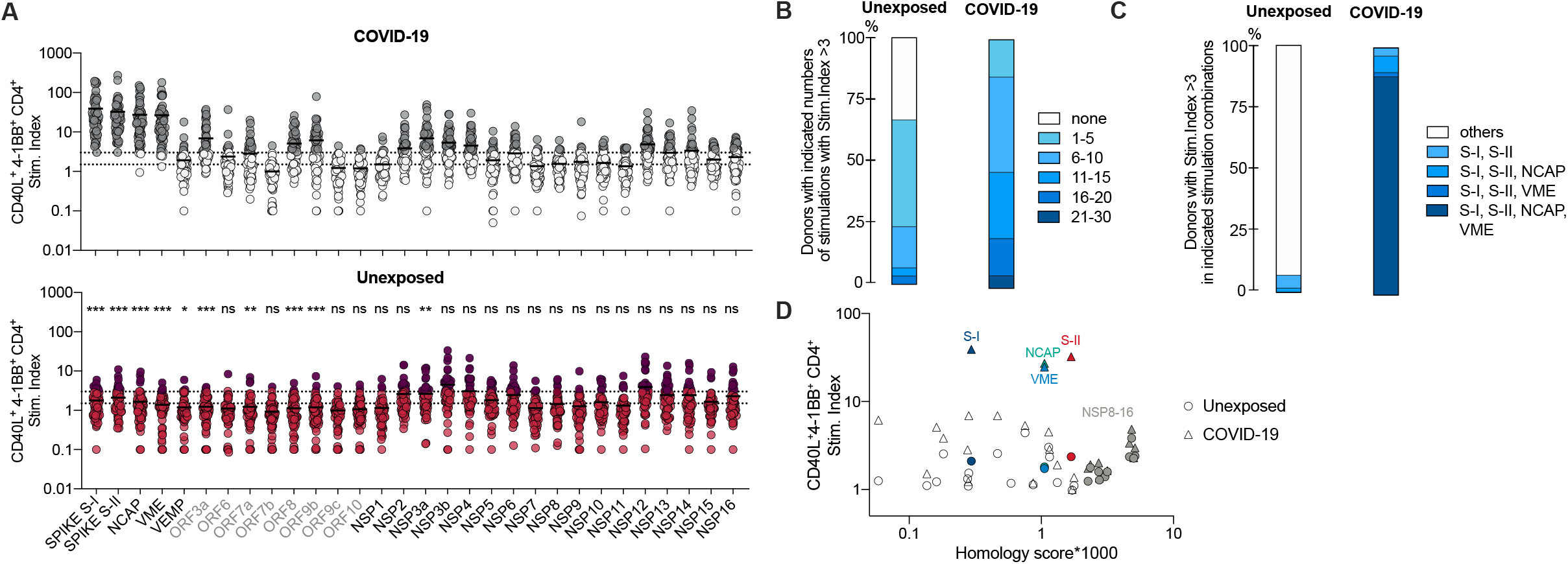
CD4^+^ T cell cross-reactivity against the SARS-CoV-2 orfeome. **(A)** Stimulation index (stim. index) of reactive CD40L^+^ 4-1BB^+^ CD4^+^ T cells among PBMCs stimulated with indicated SARS-CoV-2-orfeome peptide pools. Grey labels below indicate proteins exclusive for SARS-CoV-2 (not shared with HCoVs). Grey (COVID-19) or dark red (Unexposed) dots mark donors with an SI ≥ 3. n = 60 unexposed, n = 59 COVID-19 convalescents. Dotted lines indicate stim. index of 1.5 and 3. *p<0.05, **p<0.01, ***p<0.001 and ns = not significant for p>0.05 (unpaired student’s *t*-test). Results for statistical significance are shown above Unexposed plot. **(B)** Proportion of individuals with indicated number of SARS-CoV-2-orfeome peptide pool stimulations with a stim. index ≥ 3. **(C)** Proportion of individuals with stim. index ≥3 for each stimulation in each indicated stimulation combination. **(D)** To determine the probability of homologous epitopes, a homology score for the comparison of a SARS-CoV-2 antigen with all 4 endemic coronaviruses 229E, NL63, OC43 and HKU1 (isolates N1, N2 N5) is plotted against the mean stim. index of 60 unexposed donors (circles) and 59 COVID-19 convalescents (triangles). For the calculation of the homology score all theoretical possible 9mers in each peptide of a respective PepMix were scored against all 9mers of the complete proteome of all other HCoVs using a PAM30 substitution matrix. The homology score is the percentage of comparisons between the 9mers from the PepMix and all 9mers from the HCoVs with a score above 30.

## SARS-CoV-2 spike S-II-cross-reactive T cells decrease with age

A striking feature of SARS-CoV-2 infection is the high correlation of age with disease severity. Immunologically, aging is associated with a lack of newly generated T cells and clonal expansion of few clones as a result of persistent infections, which limits the breadth and quality of T cell responsiveness (*19*). In order to assess the effect of age on SARS-CoV-2-(cross)-reactive T cell immunity we first examined the breadth of SARS-CoV-2 spike-specific CD4^+^ T cell responses in 568 unexposed individuals and 174 COVID-19 convalescents from different age groups (Fig. 2A, Table 1). COVID-19 convalescents displayed a statistically significant age-associated increase in spike S-I reactive T cells that correlated with higher disease severity in the older cohorts (Table 1). However, in line with our previous findings (3) in unexposed individuals, T cell cross-reactivity to the N-terminal portion of spike (S-I) was rare, close to the limit of detection, and remained at low levels with increasing age, whereas reactivity to the C-terminal portion of spike (S-II) was more frequent and higher but decreased with increasing age. T cells reacting to a peptide pool representing a mixture of selected T cell epitopes from common pathogens (CEFX pool), by contrast, remained stable with age (Fig. 2A). Thus, elderly individuals exhibited decreased cross-reactivity immunity to the more HCoV-homologous spike S-II portion.

**Fig. 2:**
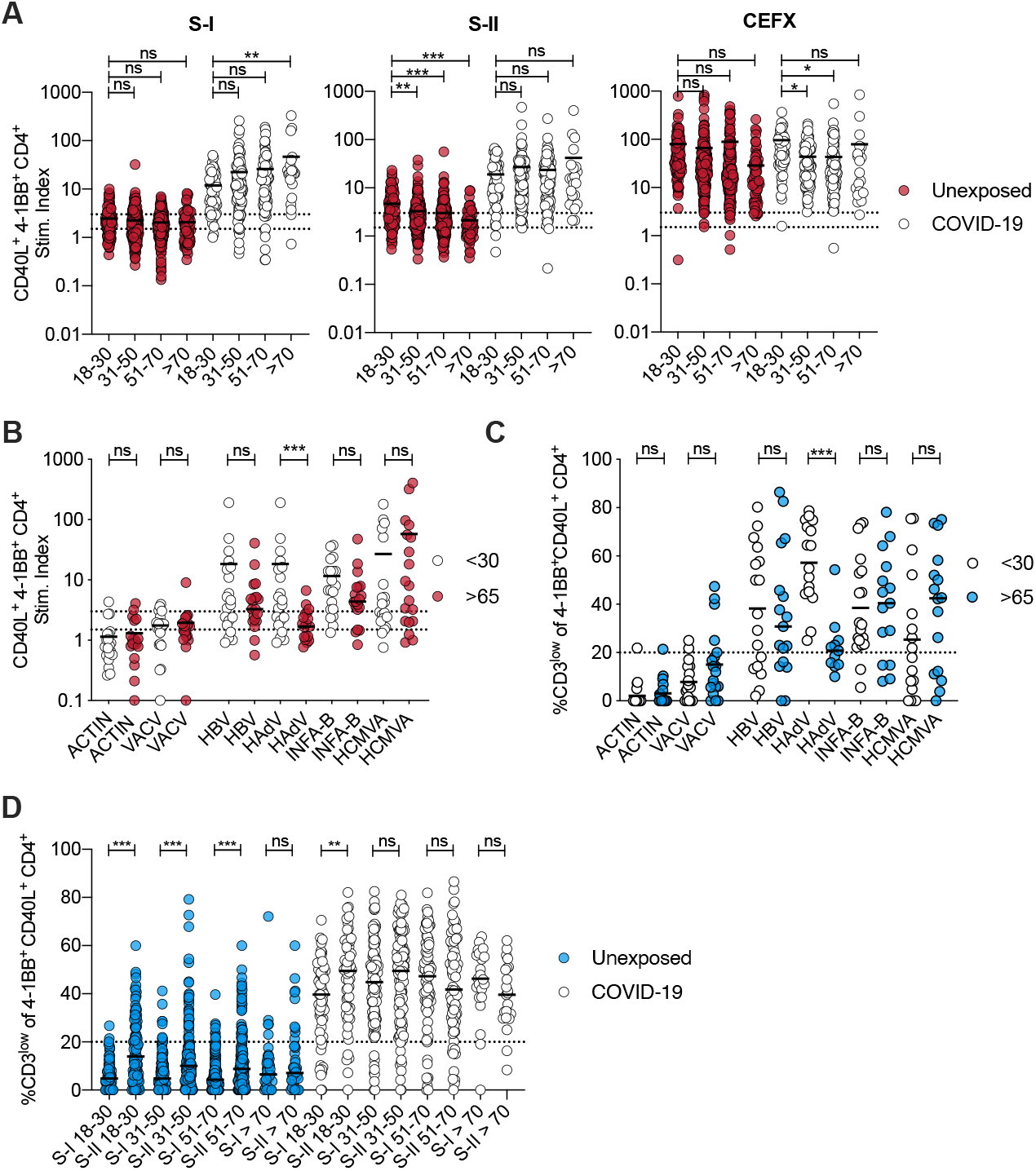
Magnitude of SARS-CoV-2-cross-reactivity decreases with age. **(A)** Stim. index of CD40L^+^ 41BB^+^ CD4^+^ T cells after stimulation of PBMCs with SARS-CoV-2 S-I, SARS-CoV-2 S-II, or CEFX (CMV, EBV, Flu and other common pathogens epitope pool) in *n* = 568 unexposed donors and *n* = 174 COVID-19 convalescents divided into age groups. **(B, C)** Stim. index of CD40L^+^ 4-1BB^+^ CD4^+^ T cells and (C) frequencies of CD3^low^ cells among CD40L^+^ 4-1BB^+^ CD4^+^ T cells after PBMC stimulation with indicated peptide pools. CD3^low^ frequencies are shown for antigen responding donors (stim. index ≥1.5) except for ACTIN and VACV where CD3^low^ frequencies are shown for all CD40L^+^ 4-1BB^+^ T cells as low control. Dotted lines indicate stim. index of 1.5 and 3 (B) or 20% (C). **(D)** Frequency of CD3^low^ cells among S-I or S-II-reactive CD40L^+^ 4-1BB^+^ CD4^+^ T cells in different age groups after stimulation of PBMC from unexposed and COVID-19 convalescents. CD3^low^ frequencies are shown for T cell responses with a stim. index ≥1.5. A: *p<0.05, **p<0.01, ***p<0.001, and ns for p>0.05, One-way-ANOVA with Dunnett correction. B, C, D: *p<0.05, **p<0.01, ***p<0.001, and ns for p>0.05 (Student’s *t*-test).

## Low CD3 surface expression identifies SARS-CoV-2 reactive T cells with high functional avidity ex vivo

To assess the quality of the spike-(cross-)reactive T cell response in terms of functional T cell avidity, we examined the level of CD3 surface expression in CD40L^+^ 4-1BB^+^ CD4^+^ T cells induced upon short-term *in vitro* stimulation. Strong T cell receptor (TCR) activation, characteristic of T cells with high TCR avidity, blocks recycling of the TCR:CD3 complex and hence is detectable by reduction in CD3 surface expression (*20*), hereafter referred to as high functional avidity. Thus, cognate cross-reactivity with higher probability of high functional avidity should be distinguishable from non-cognate cross-reactivity with higher probability of low functional avidity by analyzing the frequency of CD3^low^ T cells among CD4^+^ T cells activated via their TCR. Accordingly, when stimulated with the peptide pools of the autoantigen ACTIN or vaccinia virus (VACV, unvaccinated donors, Fig. 2B), the few reactive T cells exhibited only low non-cognate reactivity, characterized by low antigen avidity with CD3^low^ frequencies below 20% (Fig. 2C, Fig. S2). By contrast, T cell responses to the hepatitis B vaccine as well as to recurrent infections (Influenza A, Adenovirus and CMV, Fig. 2B) were characterized by high frequencies of reactive T cells and high frequencies of high avidity CD3^low^ cells (Fig. 2C). After stimulations with spike S-I/S-II peptide pools, COVID-19 convalescents showed high frequencies of spike S-I and S-II-activated CD4^+^ T cells that largely lacked CD3 expression (Fig. 2D). In unexposed individuals, the frequency of CD3^low^ cells among spike S-I and S-II-activated CD4^+^ T cells was markedly lower, but significantly higher frequencies of CD3^low^ cells were present among S-II-than among S-I-activated CD4^+^ T cells across all age groups (Fig. 2D). These results are consistent with the high homology of the C-terminal S-II portion of SARS-CoV-2 spike to the analogous S-II portion of HCoV spike. These results indicate that spike S-II (cross)-reactive CD4^+^ T cells comprise cells with high functional avidity.

## HCoV spike-reactive high functional avidity CD4^+^ T cells decreases with age

We hypothesized that previous HCoV exposures induce cognate cross-reactive CD4^+^ T cells. Therefore, we next characterized CD4^+^ T cell immunity to HCoV spike in unexposed individuals and COVID-19 convalescents, again grouped by age. HCoV-S-I and S-II-reactive CD4^+^ T cells were more readily detectable than SARS-CoV-2 spike-specific T-cells and found in 80% (S-I) and 98% (S-II) of SARS-CoV-2 unexposed individuals, respectively (Fig. 3A). However, their frequency significantly decreased with age, and in this instance this reduction was also detectable for the S-I-reactive T cells. Aggregated as a group, SARS-CoV-2 infection overall did not result in higher HCoV-S-I-/S-II-reactive T cell frequencies in COVID-19 convalescents compared to unexposed individuals. HCoV-reactive T cells were also found to decrease as a function of age. We also examined the functional avidities of HCoV-reactive CD4^+^ T cells (Fig. 3B). Among HCoV spike S-I- and S-II-reactive CD4^+^ T cells, high frequencies of CD3^low^ T cells were found, which again significantly decreased with increasing age. These results suggest a high degree of HCoV exposure of the population, leading to equally widespread cross-immunity to SARS-CoV-2 spike. HCoV-reactive CD4^+^ T cells frequently comprised cells with high functional avidity but significantly decreased with age.

**Fig. 3:**
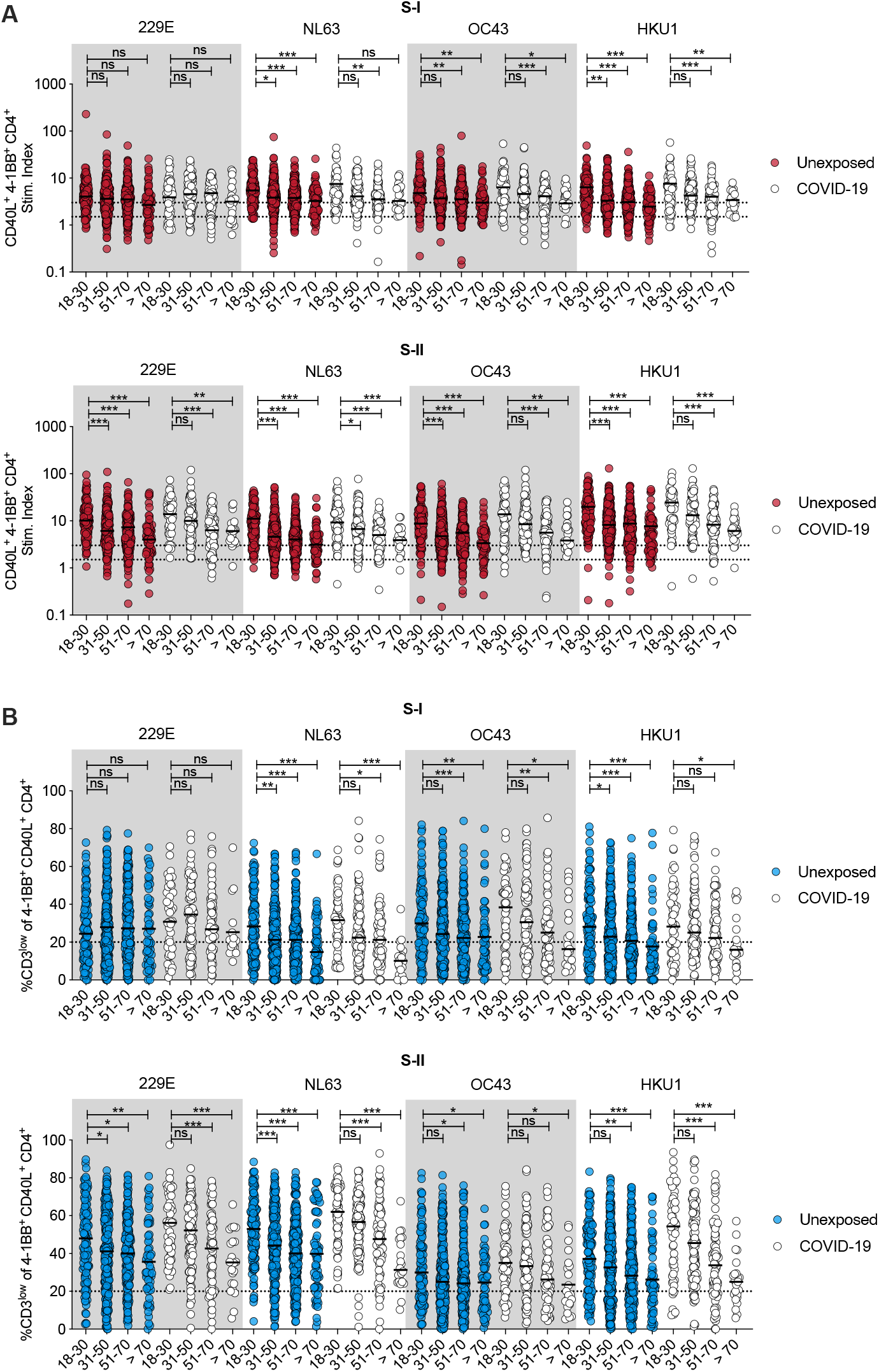
High functional avidity T cells to spike S-II from HCoVs decrease with age. **(A)** Stim. index of CD40L^+^ 4-1BB^+^ CD4^+^ T cells in unexposed (*n* = 568) as well as COVID-19 convalescents (*n* = 174) after PBMC stimulation with HCoV (229E, NL63, OC43, HKU1) spike glycoprotein S-I or S-II peptide pools. **(B)** Frequency of CD3^low^ cells in CD40L^+^ 4-1BB^+^ CD4^+^ T cells from unexposed and COVID-19 convalescents in age groups after stimulation with S-I and S-II spike glycoprotein pools of HCoVs. CD3^low^ frequencies are shown for T cell responses with a stim. index ≥1.5. A, B: *p<0.05, **p<0.01, ***p<0.001 and ns for p>0.05 (One-way-ANOVA with Dunnett correction).

## Identification of the immunodominant peptide “iCope” recognized by SARS-CoV-2 spike glycoprotein S-II-cross-reactive CD4^+^ T cells

All SARS-CoV-2-cross-reactive unexposed donors showed a response against at least 2 (S-I) or 3 (S-II) HCoVs, suggesting that repeated infection with different HCoVs either establishes a detectable prominent SARS-CoV-2 cross-reactive T cell pool and/or specific T cells are directed against highly homologous sequences shared by multiple HCoVs and SARS-CoV-2 (Fig. 4A). We examined next, whether HCoV spike glycoprotein-specific T cells directly cross-react to SARS-CoV-2 spike glycoprotein. Therefore, CD40L^+^ 4-1BB^+^ OC43 S-I or S-II-reactive CD4^+^ T cells were isolated, expanded *in vitro* and restimulated with autologous APC in the presence of OC43- or SARS-CoV-2 spike pool S-I and S-II, respectively. 6 out of 18 OC43 S-II specific short-term T cell lines displayed cross-reactivity against SARS-CoV-2 S-II (Fig. 4B) whereas OC43 S-I specific T cell lines lacked cross-reactivity against SARS-CoV-2 S-I. Next, we aimed to identify the underlying SARS-CoV-2 S-II cross-reactive epitopes. To this end, we isolated SARS-CoV-2 S-II-cross-reactive CD4^+^ T cells from 5 unexposed individuals at high purity, expanded the cells *in vitro*, and examined their specificity upon restimulation with peptide matrices. We identified two T-cell stimulating peptides (peptides 204 and 205) within S-II in all 5 donors (Fig. S3A). Restimulation of the expanded T cell lines with single peptides and negative control peptides validated the specificity for peptides 204 (N’-SKRSFIEDLLFNKVT-C’) and 205 (N’-FIEDLLFNKVTLADA-C’) (Fig. S3B). Only one donor responded to some of the other identified candidates (peptides 188, 189, 251; fig. S3B). Sequence alignment revealed that S-II peptides 204 and 205 were located in a region of spike characterized by high homology with HCoV (Fig. S3C). The peptides 204 and 205 largely overlapped and differed in only 3 amino acids (aa). By designing 15 aa peptides covering the intersection of both peptides we identified the fusion peptide domain 1 sequence N’-SFIEDLLFNKVTLAD-C’ (aa 816-830) as the ***i***mmunodominant ***Co***ronavirus ***p***eptide ***e***pitope hereafter referred to as „*iCope*“ (peptide 204_3, Fig. S3D). We next examined direct *ex vivo* T cell reactivity against *iCope* compared to a control peptide 284 (aa 1133-1147) and the SARS-CoV-2 spike S-II peptide pool in 48 unexposed individuals and 22 COVID-19 convalescents. Note that *iCope*-reactive CD4^+^ T cells were detected in 50% of convalescents and 20% of unexposed individuals with significantly higher frequency in the former (Fig. 4C). Antibodies to the SARS-CoV-2 spike amino acid residues 815-823 (peptide 204_-1) were previously reported in COVID-19 patients but also in unexposed individuals (*21*). By examining sera from *iCope*-T-cell-assay-responders and non-responders we detected 204_-1 binding antibodies in all individuals, however, significantly higher concentration of these antibodies was found in COVID-19 convalescents with substantially more *iCope*-reactive T cells (Fig. 4D). HLA-typing of *n* = 308 unexposed study participants revealed that among the most frequent alleles (found in >10% of individuals in the study) definite *iCope* responders (stim. index ≥ 3) compared to definite non-responders (stim. index ≤ 1.5) were more frequently positive for HLA-DPB1*02:01, HLA-DPB1*04:02 and, especially homozygous expression of HLA-DPB1*04:01 (Fig. 4E). Since HLA-DPA1*01:03 was found in 100% of the responders and 94.8% of the non-responders we investigated if combinations of HLA-DPA1*01:03 and HLA-DPB1*02:01/DPB1*04:01/DPB1*04:02 were likely to present *iCope* or fragments thereof. HLA-peptide-binding predictions (www.IEDB.org) identified excellent potential binders (Fig. S3E) which was also true for the homologous peptide of *iCope* in other HCoVs (Fig. S3F).

**Fig. 4:**
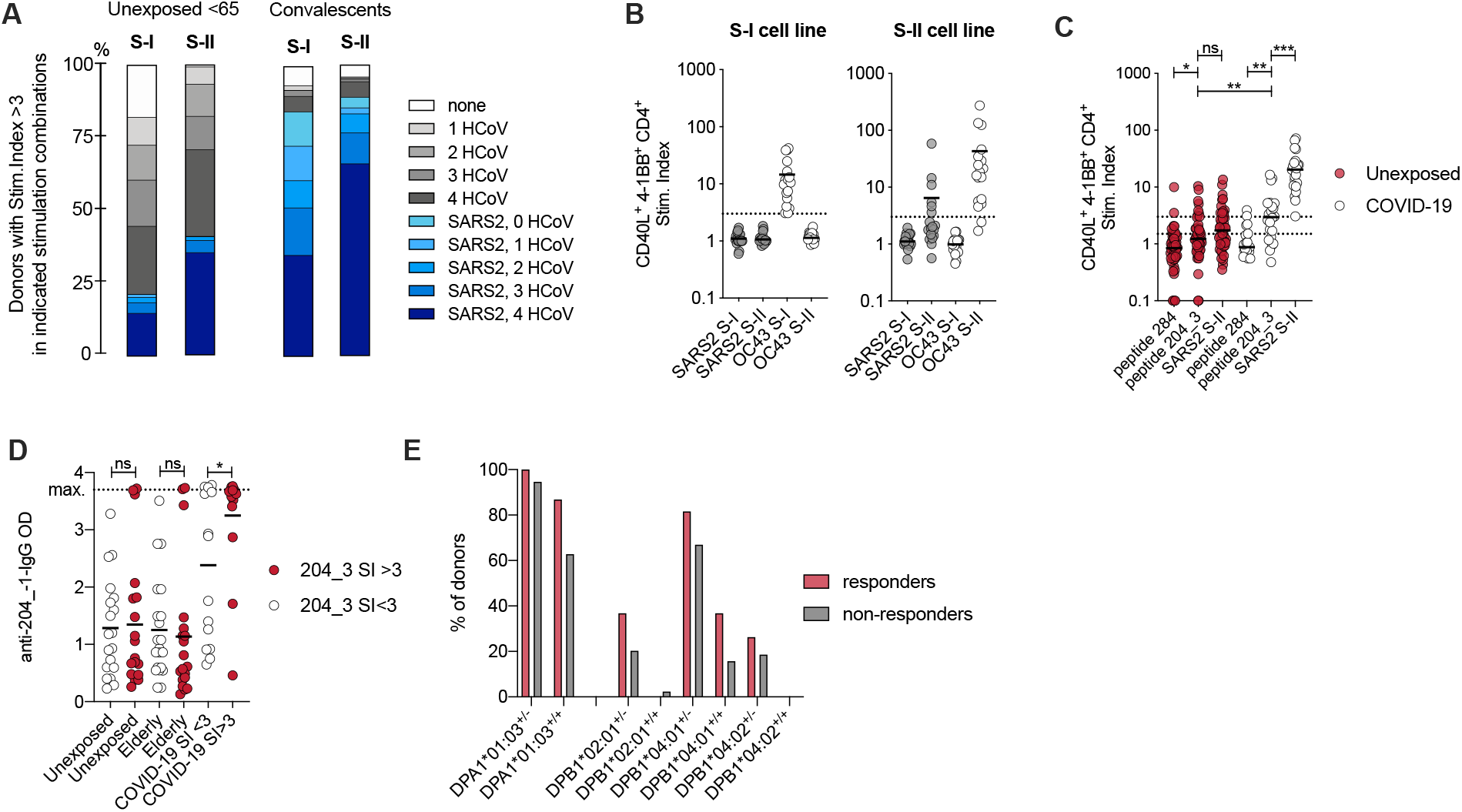
Peptide 204_3 (*iCope*) constitutes an immunodominant epitope of SARS-CoV-2 T cell (cross)-reactivity. **(A)** Proportions of unexposed individuals aged below 65 (*n* = 491) and COVID-19 convalescents (*n* = 174) with S-I or S-II specific responses against HCoV and/or SARS-CoV-2 with a stim. index ≥ 3. **(B)** Short-term T cell lines derived from OC43 S-I and S-II-reactive primary T cells were restimulated with autologous APC in the presence of OC43 or SARS-CoV-2 spike glycoprotein pool S-I and S-II. Stim. index of CD40L^+^ 4-1BB^+^ CD4^+^ T cells after re-stimulation is shown. Dotted line indicates stim. index of 3. **(C)** Stim. index of CD40L^+^ 4-1BB^+^ CD4^+^ T cells from unexposed or COVID-19 convalescents after stimulation with the single peptide 204_3, the control single peptide 284 or the S-II peptide pool. **(D)** Anti-204_-1-peptide IgG ELISA. The ELISA plates were coated with a 18mer peptide overlapping 11 amino acids with the 204_3 peptide. Serum from unexposed and elderly (>65) individuals as well as COVID-19 convalescents were used at 1:100 dilution. Optical density (OD) of detected IgG antibodies is shown. **(E)** Distribution of common HLA alleles in definite *iCope* responders (stim. index ≥ 3) and definite non-responders (stim. index ≤ 1.5), *n* = 308, ^+/+^ = homozygous, ^+/-^ = heterozygous. C, D: *p<0.05, **p<0.01, ***p<0.001 and ns for p>0.05 (Student’s *t*-test).

## Pre-existing SARS-CoV-2 S-II-cross-reactive T cells are recruited into the immune response to primary SARS-CoV-2 infection

One of the still outstanding questions is to what extent and how SARS-CoV-2-cross-reactive T cells influence the disease course of primary SARS-CoV-2 infection. Therefore, the Charité Corona Cross study recruited 760 healthy unexposed individuals from July 2020 to February 2021 with the objective to recruit them again in case of diagnosed primary SARS-CoV-2 infection. Baseline immunity to spike S-I and S-II of SARS-CoV-2 and all 4 HCoVs was determined in each subject at the first visit. Study participants informed us in the event of symptoms suggestive of SARS-CoV-2 infection, after which multiplex RT-PCR screening for SARS-CoV-2 and all 4 HCoVs was performed. We aimed to re-sample patients on day (d) 7, d14, d21 and >d35 after the assumed infection day by estimating the infection day to be three days before symptom onset. 17 cases of acute primary SARS-CoV-2 infection were identified (Table 2, Fig. 5). All 17 patients showed mild COVID-19 disease progression and immediately detectable viral titers (Fig. S4). Robust CD4^+^ T cell responses specific of SARS-CoV-2 spike S-I and S-II were detected and the proportions of HLADR^+^ CD38^+^ cells among CD40L^+^ 4-1BB^+^ CD4^+^ T cells significantly increased on follow up time points 1 and 2 (3-16 days) indicating their in *vivo* activation (Fig. 5A, B). High functional avidity CD3^low^ cells substantially increased during acute primary SARS-CoV-2 infection and remained at a high level when the infection had resolved (Fig. 5C). Individuals who already had spike S-II-cross-reactive CD4^+^ T cells with a stimulation index above 3 at baseline showed superior high functional avidities throughout the initiation of the T cell response (Fig. 5D). *iCope*-reactive T cells increased in frequency and in functional avidity in 10 of 17 donors after infection (Fig. 5E, F). Notably, IgG antibodies against the 204_-1 peptide were boosted as early as 3-9 days (follow-up 1) after the presumed infection (Fig. 5G). The appearance kinetics and levels of anti-S1-IgG serum antibodies varied widely, being first detectable at follow-up time point 2 and peaking after day 20 in most subjects (Fig. 5H). Anti-S1-IgG ratios at late time points positively correlated with S-II-but not S-I-cross-reactive T cell levels at d0 suggesting that pre-existing cross-reactive CD4^+^ T cells enhanced SARS-CoV-2-specific humoral immunity (Fig. 5I, left). Moreover, the titer of neutralizing antibodies also positively correlated with S-II-but not S-I-cross-reactive CD4^+^ T cells at baseline, indicating a role of cross-reactive CD4^+^ T cells in protection (Fig. 5I, middle and right). Finally, the frequency of pre-existing HCoV-reactive CD4^+^ T cells increased in almost all individuals shortly after primary SARS-CoV2 infection (Fig. 5J). There was also a concomitant increase in the frequency of CD3^low^ cells (Fig. 5K) and HLADR^+^ CD38^+^ cells (Fig. 5L) among HCoV-reactive T cells, demonstrating that pre-existing HCoV-reactive cellular immunity became activated and expanded during primary SARS-CoV-2 infection. Overall, these results indicate that pre-existing SARS-CoV-2 S-II-cross-reactive T cells were recruited into primary SARS-CoV-2 immune responses in healthy unexposed individuals. The quantity and functional avidity of pre-existing cross-reactive cellular immunity corresponded to the quality and magnitude of specific cellular and humoral anti-SARS-CoV-2 responses and thus may efficiently support mild COVID-19 disease courses.

**Table 2:**
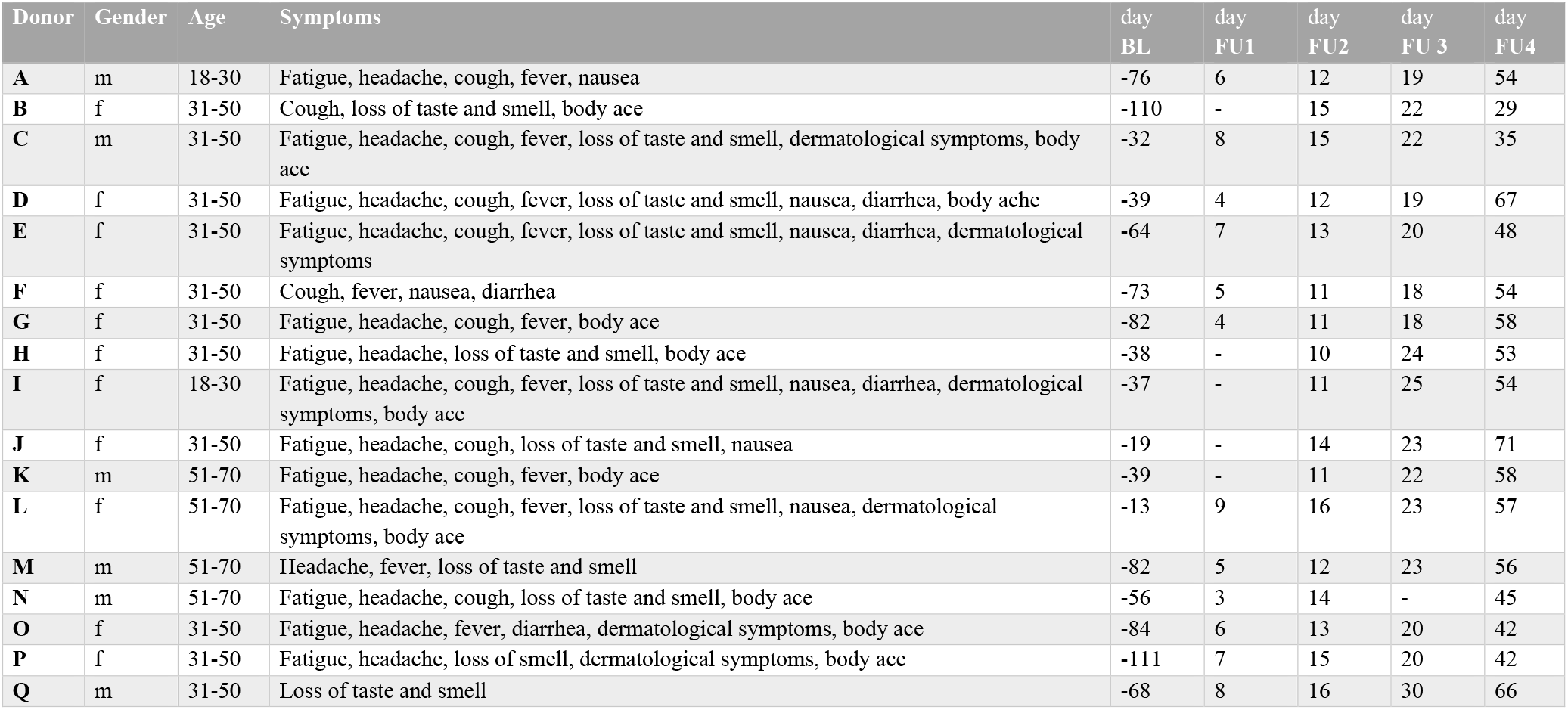
Characteristics of the follow-up donors. BL = baseline, FU = follow-up. None of the donors required hospitalization.

**Fig. 5:**
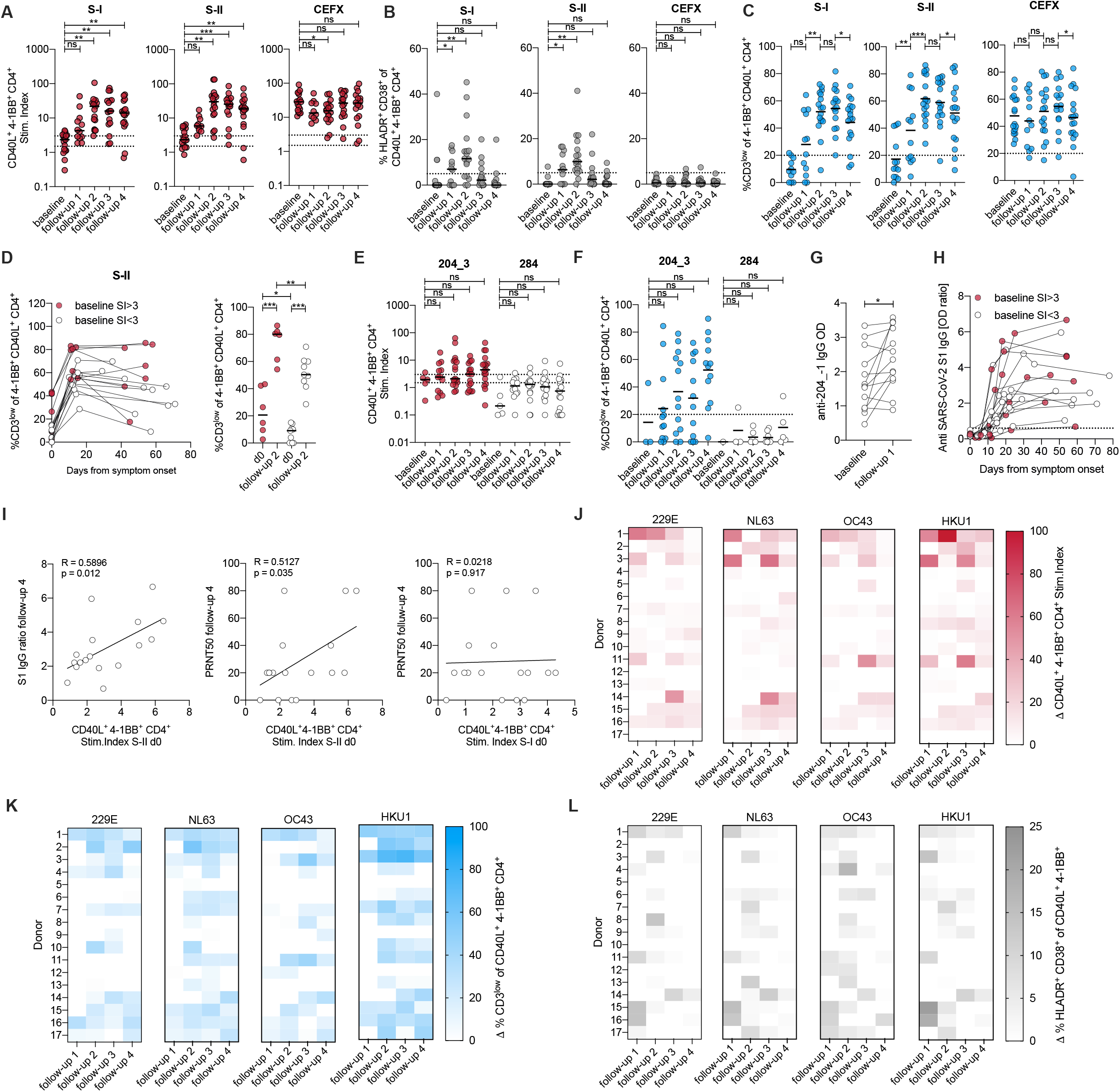
HCoV-specific SARS-CoV-2-cross-reactive T cells are recruited into the primary SARS-CoV-2 infection response. **(A-C)** Stim. index of CD40L^+^ 4-1BB^+^ CD4^+^ T cells (A), frequencies of HLADR^+^ CD38^+^ (B), and frequencies of CD3^low^ (C) in CD40L^+^ 4-1BB^+^ CD4^+^ T cells after stimulation with SARS-CoV-2 S-I, S-II and CEFX peptide pools of donors prior (baseline) to and at 4 different time points after SARS-CoV-2 infection. CD3^low^ frequencies are shown for T cell responses with a stim. index ≥1.5. **(D)** Overview of changes in CD3^low^ frequency in CD40L^+^ 4-1BB^+^ CD4^+^ T cells between baseline, follow-up measurement time point 2 (between d10-d16 after symptom onset) and follow-up measurement time point 4 (between d29-d71) (left plot) and statistics (right plot) for baseline and follow-up measurement time point 2 in cross-reactive donors (baseline stim. index ≥ 3, red circles) and non-cross-reactive donors (baseline stim. index ≤ 3, white circles). **(E)** Stim. index of CD40L^+^ 4-1BB^+^ CD4^+^ T cells and **(F)** frequency of CD3^low^ of CD40L^+^ 4-1BB^+^ CD4^+^ T cells after stimulation with peptide 204_3 (*iCope*) or control peptide 284. CD3^low^ frequencies are shown for T cell responses with a stim. index ≥1.5. **(G)** Anti-204_- 1-peptide IgG ELISA. The ELISA plates were coated with an 18-mer peptide overlapping 11 amino acids with the 204_3 peptide. Sera from baseline and 7 days after symptom onset were used at a 1:100 dilution. Optical density (OD) of detected IgG antibodies is shown. **(H)** Kinetics of anti-S1-IgG antibody ELISA OD ratios in cross-reactive donors (baseline stim. index ≥ 3, red circles) and non-cross-reactive donors (baseline stim. index ≤ 3, white circles). **(I)** Correlation of anti-S1-IgG antibody ELISA OD ratio at last sampling time point FU4 (29-71 days after symptom onset, cf. Table 2) to the frequency of CD40L^+^ 4-1BB^+^ CD4^+^ upon S-II stimulation at d0 (baseline, left plot), and correlation of neutralizing antibody titers PRNT50 at last sampling time point FU 4 to frequency of CD40L^+^ 4-1BB^+^ CD4^+^ upon stimulation with S-II (middle plot) or to stimulation with S-I at d0 (baseline, right plot). **(J, K, L)** Heat maps with the delta (Δ) of stim. index (J), frequency of CD3^low^ (K) and frequency of HLADR^+^ CD38^+^ cells (L) of CD40L^+^ 4-1BB^+^ CD4^+^ T cells after stimulation with S-II pools of indicated HCoVs. Delta is the change of the parameter at the given time point relative to baseline, i.e., white depicts no increase. A, C, D, F, G: *p<0.05, **p<0.01, ***p<0.001 and ns for p>0.05 (Paired Student’s *t*-test). B: *p<0.05, **p<0.01, ***p<0.001 and ns for p>0.05 (repeated measure one-way-ANOVA with Dunnett correction). E: ns for p>0.05 (paired Student’s *t*-test). I: Pearson correlation.

## BNT162b2 vaccination re-activates pre-existing SARS-CoV-2 spike S-II-cross-reactive T cells

Finally, we investigated how pre-existing SARS-CoV-2 S-II-cross-reactive T cells in healthy unexposed individuals influenced the course of BNT162b2 COVID-19 spike mRNA vaccine responses. We monitored baseline- and follow up humoral- and T cell responses against SARS-CoV-2- and HCoV spike glycoproteins in 31 healthy adults who underwent primary and booster vaccination with BNT162b2 at d0 and d21. At day 21, 30 of 31 donors had seroconverted for anti-S1-IgG and all donors had seroconverted for anti-S1-IgA (Fig. 6A) with booster vaccination further increasing the antibody levels. BNT162b2 vaccination induced robust S-I and S-II CD4^+^ T cell responses in all individuals after primary vaccination which were again slightly enhanced by booster vaccination (Fig. 6B). By contrast, the functional avidity of CD40L^+^ 4-1BB^+^ CD4^+^ T cells was again substantially increased by booster vaccination (Fig. 6D). Moreover, at day 14 all but three donors had high frequencies of HLADR^+^ CD38^+^ cells among S-I- and S-II-reactive CD4^+^ T cells indicating their recent *in vivo* activation (Fig. 6E). Of note, kinetics of S-I- and S-II-reactive T cells differed in that S-II-reactive T cells showed a sharp increase from baseline to 7 days after vaccination but not thereafter, whereas S-I-reactive T cells again showed a marked increase from day 7 to day 14 (Fig. 6B, C), which is indicative of secondary response kinetics of the former and primary response kinetics of the latter (*22*). Frequencies of HCoV S-II-reactive T cells were significantly increased 7 days after primary vaccination (Fig. 6F), which was in accordance with increased frequencies of HCoV S-II-reactive HLADR^+^ CD38^+^ T cells (Fig. 6G), demonstrating that cognate cross-reactive T cells were activated in response to SARS-CoV-2 spike-specific vaccination. All but 2 of 31 donors (94%) responded with high functional avidity T cells to *iCope* at day 7 and day 14 (Fig. 6H, I). We observed a correlation between the *iCope-reactive* T cell response and the S-II-reactive T cell response at d0 that was even more pronounced in the early stage of the immune response on day 7 emphasizing the importance of *iCope* in the anti-SARS-CoV-2 cellular immune response (Fig. 6K). *iCope*-specific T cells initially contributed to up to 100% of the CD40L^+^ 4-1BB^+^ cells in S-II stimulations but decreased in proportion as other specificities increased during the course of the immune response (Fig. 6L). Finally, comparable to the cellular response, a humoral response to the *iCope*-overlapping sequence 204_-1 was detectable upon vaccination as early as 7 days after primary vaccination (Fig. 6M) supporting the notion of secondary response kinetics (*23*) for pre-existing cross-reactive immunity.

**Fig. 6:**
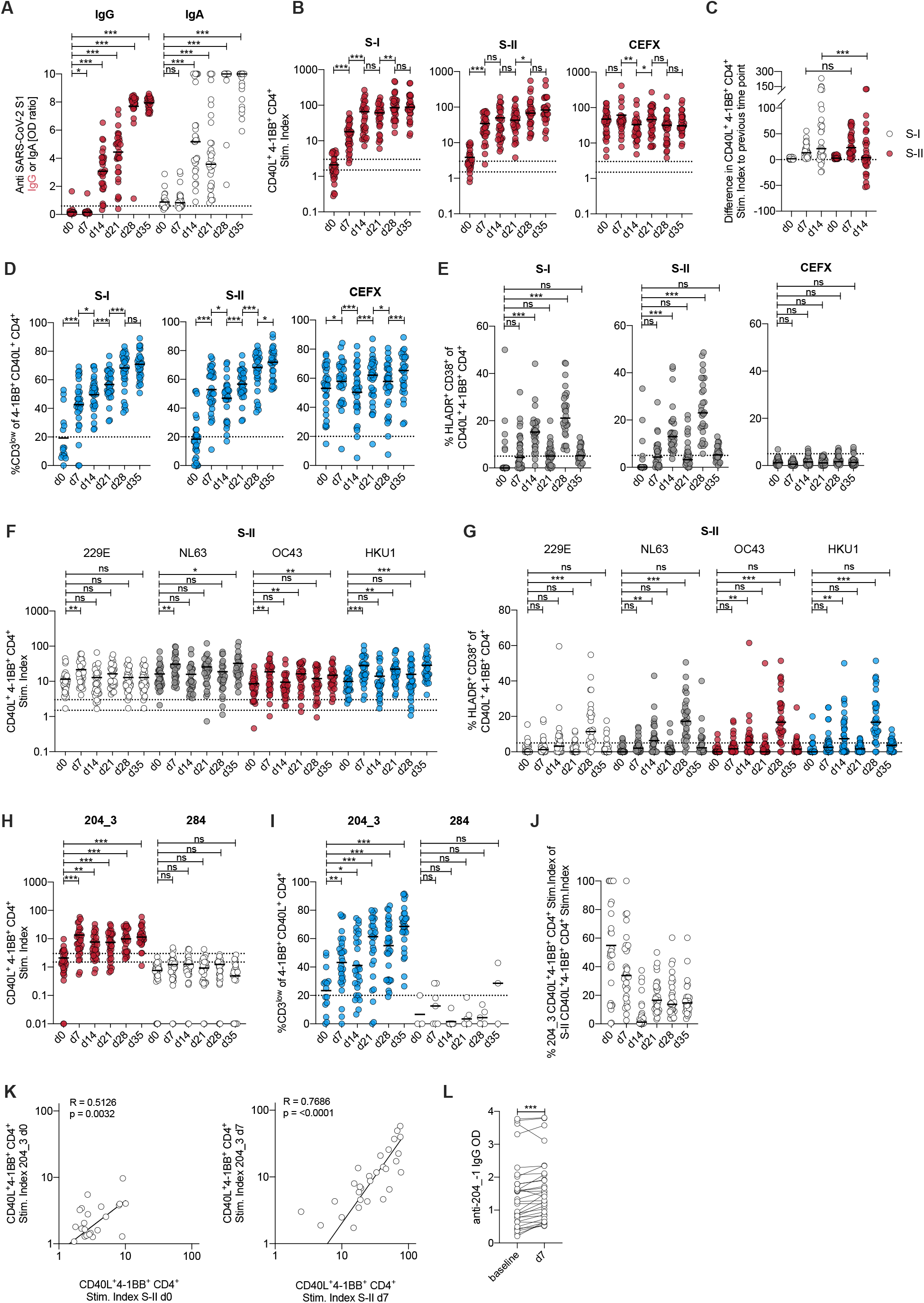
HCoV-specific SARS-CoV-2-cross-reactive T cells are recruited into the BNT162b2 vaccine response. **(A)** Serum IgG and IgA SARS-CoV-2 S1 ELSIA OD ratio at baseline (d0) and d7, d14, and d21 after primary immunization with the BNT162b2 COVID-19 vaccine, as well as d7 (d28) and d14 (d35) after secondary vaccination. Secondary vaccination occurred on day 21. IgA max. value set as 10. **(B)** Stimulation index of CD40L^+^ 4-1BB^+^ CD4^+^ T cells after stimulation with S-I, S-II and CEFX. **(C)** Difference of stim. index relative to the previous time point measured. **(D)** Frequency of CD3^low^ of CD40L^+^ 4-1BB^+^ CD4^+^ T cells after stimulation with S-I, S-II and CEFX. CD3^low^ frequencies are shown for T cell responses with a stim. index ≥1.5. **(E)** Frequencies of HLADR^+^ CD38^+^ of CD40L^+^ 4-1BB^+^ CD4^+^ T cells after stimulation with S-I, S-II and CEFX. **(F, G)** Stim. index (E) and frequency of HLADR^+^ CD38^+^ (F) of CD40L^+^ 4-1BB^+^ CD4^+^ T cells after stimulation with HCoV S-II peptide pools at baseline and indicated days after primary vaccination. **(H, I)** Stim. index (G) and frequencies of CD3^low^ (H) of CD40L^+^ 4-1BB^+^ CD4^+^ T cells after stimulation with peptide 204_3 (*iCope*) and control peptide 284. CD3^low^ frequencies are shown for T cell responses with a stim. index ≥1.5. **(J)** Ratio of 204_3-reactive T cells to SARS-CoV-2 S-II-reactive T cells. **(K)** Correlation of stimulation with single peptide 204_3 to stimulation with SARS-CoV-2 S-II peptide pool at d0 or d7, respectively for vaccinees. **(L)** Anti-204_-1-peptide IgG ELISA. The ELISA plates were coated with an 18-mer peptide overlapping 11 amino acids with the 204_3 peptide. Sera from before and 7 days after primary vaccination were used at a 1:100 dilution. Optical density (OD) of detected IgG antibodies is shown. A, E, F, G, H, I: *p<0.05, **p<0.01, ***p<0.001 and ns for p>0.05 (repeated measure one-way-ANOVA with Dunnett correction). B, C, L: *p<0.05, **p<0.01, ***p<0.001 and ns for p>0.05 (Paired Student’s *t*-test). D: *p<0.05, **p<0.01, ***p<0.001 and ns for p>0.05 (Student’s *t*-test). K: Pearson correlation.

## Discussion

A better understanding of how cross-reactive immunity influences COVID-19 disease severity and virus clearance is central to containment efforts as well as to the evaluation of the most efficient vaccination strategies. Non-cognate cross-reactivity has been reported but plays a minor role compared to HCoV-mediated cognate cross-reactivity (16,23). However, the clinical relevance of pre-existing cognate cross-immunity to SARS-CoV-2 has been a subject of much debate. A recent HCoV infection has been associated with less severe COVID-19 (*17*). Interestingly, more than 90% of the population is HCoV seropositive, thus a large proportion of the population might have some underlying humoral immunity (24, 25). However, in pre-pandemic sera from PCR-validated HCoV-positive individuals, neutralizing antibodies against all HCoVs but no neutralizing antibodies to SARS-CoV-2 were found (*24*). In a further report, only low spike-specific cross-reactive antibody activity was detected in 5 of 34 donors with recent HCoV infection and in 1 of 31 donors without recent HCoV infection, indicating low level and fast decay of humoral cross-immunity (12). Finally, infection with SARS-CoV-2 increased the prevalence of antibodies against seasonal HCoVs which, however, did not provide protection against SARS-CoV-2, thus drawing attention to the role of cross-reactive cellular immunity (*9, 24, 25*).

Recently, cross-reactive T cells in unexposed individuals for several SARS-CoV-2 antigens utilizing predicted epitopes (*4, 5*) or megapools of selected peptides were identified (*8, 26*). Although the 21kb long ORF1a/b accounts for the majority of the total 29.8 kb coding sequence of SARS-CoV-2, the totality of T cell responses to ORF1a/b encoded proteins in COVID-19 convalescents compared to cross-reactive T-cell responses in healthy unexposed donors had not been previously analyzed. Our work revealed significant cross-reactivity regarding ORF1a/b encoded proteins but also that most of the anti-SARS-CoV-2 reactivity was directed against spike, NCAP and VME protein. We further demonstrated that the magnitude and quality of SARS-CoV-2 cross-reactivity and HCoV-reactivity declined with age. This failure of the aging immune system to maintain HCoV-induced SARS-CoV-2 cross-reactive T cells might explain, to an extent, the significant increase in life-threatening COVID-19 disease in the older population. Our results demonstrate not only that HCoV-specific, cross-reactive T cells contributed to SARS-CoV-2 immune responses upon infection and vaccination, but also, that cognate cross-reactivity correlates with a slightly faster high quality cellular as well as enhanced humoral response. Mechanistic insights into this observation can be derived from prime-boost experiments in mice. Here, sequential administration of two antigens sharing the same carrier induces pre-existing T-cell help leading to more efficient B-cell recruitment in secondary immunization (*27*). Further studies in mice showed that T cell signals early in the immune response can be limiting and that after viral infection, the presence of cognate T cell help promotes germinal center formation which is required for high affinity antibody generation (*27*–*29*). As early induction of SARS-CoV-2 T cell reactivity has been associated with rapid viral clearance and mild disease (*30*), cross-reactive T cells that enhance the immune response to SARS-CoV-2 may be a correlate of immune protection, as recently discussed (*31, 32*).

Upon BNT162b2 vaccination we observed immune responses that exceeded the response to SARS-CoV-2 infection in terms of T cell and antibody levels. Responses to S-II, unlike responses to the non-cross-reactive S-I, displayed a kinetics profile reminiscent of a secondary rather than a primary response (*22, 23*). These observations may provide an explanation for the results of current large on-going studies showing protection levels against SARS-CoV-2 infection in excess of 75% as early as 15–28 days after primary vaccination with BNT162b2 (*33*). In addition, just one dose of the BNT162b2 or the Astra Zeneca ChAdOx1 vaccine reduced the risk of hospitalization by 85% and 94%, respectively, on days 28-34 post primary vaccination, indicating markedly high efficiency (*34*). Interestingly, a single shot vaccination protocol based on A26 adenovirus encoded, modified spike protein from Johnson & Johnson with a reported vaccine efficiency of 66% was recently approved by the FDA and the EMA (Janssen Ad26.COV2.S (COVID-19) Vaccine VRBPAC Briefing document, FDA). Our data supports the notion that cross-immunity induced by HCoV infections may favor milder SARS-CoV-2 infection courses and furthermore provide an immunological explanation for the high clinical efficacy of SARS-CoV-2 vaccines after just one single dose. The known abundance of HCoV infections would imply that subjects with no or reduced immunity to HCoVs would be at a higher risk of failing to control early virus replication and, as a result, provide conditions favoring viral dissemination. In unexposed subjects with sufficient levels of HCoV reactivity and thus pre-existing cross-reactivity against SARS-CoV-2, a single dose of a spike protein based vaccine may provide efficient protection. However, in older people with waning HCoV reactivity and SARS-CoV-2 cross-reactivity, repeated vaccinations may be critical (*35*).

The immunodominant cross-reactive peptide (*iCope*) that we identified here is located within the highly conserved spike fusion domain downstream of the S2′ cleavage site (*36*). We demonstrate that *iCope*-reactive T cells were efficiently recruited into the SARS-CoV-2 response in the majority of infected and almost all vaccinated individuals. The literature reports variable but generally high frequencies of HLA-DPA1*01:03, DPB1*02:01, DPB1*04:01, and DPB1*04:02 in different populations around the world (allelefrequencies.net). The high allele frequencies of HLA-DPA1*01:03 and, in particular, HLA-DPB1*04:01 as well as increased homozygosity among responders in our population may explain why so many individuals were able to raise a T cell response to *iCope* and its homologues in other HCoVs. Of note, IgG screening against spike and nucleocapsid specific epitopes revealed high responses in COVID-19 convalescents as well as multiple cross-reactive responses in healthy donors for a sequence containing the SARS-CoV-2 B cell epitope SFIEDLLF (*21*) present in *iCope* and overlapping 73% with *iCope*. SARS-CoV-2 infection and vaccination with BNT162b2 induced specific antibody production towards this region. Recently, it has been reported that in current vaccine trials, antibodies specific to the S2 portion of spike, in particular, may be involved in the early induction of protection (*25, 37*). *iCope* might serve as a universal conserved coronavirus target for both B cells and T cells. Enhancing the immune response to *iCope* may have protective effects without the possibility of immune evasion by HCoVs and should be a focus of future studies.

## Data Availability

All raw data is shown in the supplementary figures or will be provided upon request.

## Acknowledgements

The authors thank Stefan Kolling and the CCC associates Jasmin Schlotterbeck, Emre Baysal, Tobias Panne, Franziska Legler, Maxim Girod, Benedict Bohnen, Thao Nguyen, Janna Schmitz, Rebecca Klemencic, Lea Hintze, Neval Avinc, Philipp Resch, Ardit Maraj, Ahmed Farghaly, Philipp Schulz, Ke Du, Nickolai Matuschewski, Katharina Gutwenger, Ilias Katsianas, Zhuo Cheng, Andre Braginets, Gabriel Janach and Aleksander Bogdanski for their contributions to donor recruitment, sample processing and measurement and Franz Gutsch, Tobias Bleicker, Julia Tesch, Patricia Tscheak, Marie Luisa Schmidt and Johanna Riege for their support. We thank Ulf Klein for the critical reading of the manuscript.

## Author contributions

Conceptualization: LL, CGT, AT; Data curation: LL, JB, LH, VMC, TS; Formal Analysis: LL, JB, LH; Funding acquisition: AT, VMC, CD; Investigation: LL, LH, JB; Resources: UR, FKe, MM, CU, TS, FD, RL, SK, JRo, KG, ZU-A, MF, FKu, KS, ME, SH, AH, BS, SM, FP, MAMa, HW, SV, MAMu, CD, NL, LM-A, LES, VMC, JRö; Visualization: LL, JB, UR, FKe; Writing: LL, AT, CGT.

## Data and materials availability

All data are available in the main text or in the supplementary materials.

## Materials and Methods

### Study subjects

This study was approved by the Institutional Review board of the Charité (EA/152/20) and written informed consent was obtained from all participants included (*38*). Participants who tested positive for SARS-CoV-2 RNA in nasopharyngeal swabs by RT-qPCR were classified as convalescent donors. All donors were assessed for age, gender, BMI, comorbidities and medications (Table 1). Convalescent donors were subclassified according to their symptoms into WHO severity grades and information about hospitalization or admission to an intensive care unit (ICU) is given in Table 1. Day of infection was set as day −3 prior to reported symptom onset. Measurement day post symptom onset is indicated in the graphs or Table 1. Study participants who reported symptoms typical for a SARS-CoV-2 infection were RT-qPCR tested for virus RNA and positive donors were enrolled for follow-up measurements. Details of the follow-up cohort (age, gender, comorbidities, symptoms, measurement timepoints post symptom onset) are given in Table 2.

### Coronavirus RT-qPCR

RNA was extracted by using the MagNA Pure 96 system (Roche, Germany). The viral RNA extraction was performed using from 200 µl swab dilution (Swab suspended in 4.3 ml cobas PCR Media, Roche), eluted in 100µl. Coronavirus detection using 5µl of the RNA eluate was based on two genomic targets (E- and N gene, TIB Molbiol, Berlin, Germany) and detection of the four endemic CoVs was done in a multiplex format (TIB Molbiol, Berlin, Germany). An *in-vitro* transcribed RNA of equine arteritis virus was used as an internal RT and PCR control. Quantification of SARS-CoV-2 was done using the E-gene target and by applying calibration curves and using serial diluted photometrically quantified in-vitro transcribed RNA as described before (*39*). All RT-qPCR’s were performed using a LightCycler 480 II (Roche).

### Blood and serum sampling and PBMC isolation

Whole blood was collected in lithium heparin tubes for peripheral blood mononuclear cells (PBMC) isolation and SST™II advance (all Vacutainer^®^, BD) tubes for serology. SST™II advance tubes were centrifuged at 1000g, 10min and serum supernatant aliquots frozen at - 20°C until further use. PBMCs were isolated by gradient density centrifugation according to the manufacturer’s instructions (Leucosep tubes, Greiner; Biocoll, Bio&SELL).

### *Ex vivo* T cell stimulation

Freshly isolated PBMC were cultivated at 5×10^6^ PBMC in AB-medium containing RPMI 1640 medium (Gibco) supplemented with 10% heat inactivated AB serum (Pan Biotech), 100 U/ml penicillin (Biochrom), 0.1 mg/ml streptomycin (Biochrom). Stimulations were conducted with 11aa overlapping 15-mer PepMix™ SARS-CoV-2, HCoV-229E, HCoV-OC43, HCoV-NL63, HCoV-HKU1 spike glycoprotein peptide pool 1 or 2 (all from JPT) at concentrations of 1 µg/ml per peptide respectively. SARS-CoV-2 orfeome stimulations were performed with NCAP-1, VEMP-1, VME-1, AP3A (ORF3a), NS6, NS7A, NS7B, NS8, ORF9B, ORF10, Y14 (ORF9c), and the ORF1a/b pools: NSP01, NSP02, NSP03a, NSP03b, NSP04, NSP05, NSP06, NSP07, NSP08, NSP09, NSP10, NSP11, NSP12, NSP13, NSP14, NSP15, NSP16 (all from JPT) at concentrations of 1 µg/ml per peptide respectively. Used control peptide pools were hepatitis B virus protein LEP (HBV-LEP), adenovirus penton protein (HAdV5), influenza A Brisbane H1N1 (INFA-HABris), vaccinia virus (VACV), autoantigen actin, human cytomegalovirus protein pp65 (HCMVA), all from JPT. Single peptide stimulation was conducted with 1 µg/ml of the following peptides: 204 (N’-SKRSFIEDLLFNKVT-C’), 204_1 (N’-KRSFIEDLLFNKVTL-C’), 204_2 (N’-RSFIEDLLFNKVTLA-C’), 204_3 (N’-SFIEDLLFNKVTLAD-C’), 205 (N’-FIEDLLFNKVTLADA-C’) or the control peptide 284 (N’-VNNTVYDPLQPELDS-C’) (all from JPT). Stimulation controls were performed with equal concentrations of DMSO in PBS (unstimulated control) and 1.5 mg SEB/1.0 mg TSST1 (Sigma-Aldrich) or 1 µg/ml per peptide of CEFX Ultra SuperStim pool (JPT) (positive control). All approaches contained 1 µg/ml purified anti-CD28 (clone CD28.2, BD Biosciences). Incubation was performed at 37°C, 5% CO2 for 16 h in the presence of 10 µg/ml brefeldin A (Sigma-Aldrich) during the last 14 h. CD4^+^ T cell activation was plotted as Stimulation Index (Stim. index) = % of CD40L^+^ 4-1BB^+^ CD4^+^ T cells in stimulation / % of CD40L^+^ 4-1BB^+^ CD4^+^ T cells in unstimulated control. Dotted lines indicate SI of 1.5 (positive with uncertainty) and 3 (certainly positive). For CD3^low^ plots the threshold line is set at 20%.

### T cell enrichment and expansion

Activated cells were enriched from stimulated PBMCs by magnetic cell sorting (MACS). Cells were stimulated with indicated PepMixes in the presence of 1 µg/ml purified anti-CD28 (clone CD28.2, BD Biosciences) and 1 µg/ml purified anti-CD40 (5C3, Biolegend) for 16h followed by staining with CD40L-APC (5C8, Miltenyi) and 4-1BB-PE (4B4-1, BD). The activated cells were enriched with anti-PE MultiSort MicroBeads (Miltenyi) according to manufacturer’s instructions. After release of anti-PE beads a second enrichment was performed with anti-APC MicroBeads (Miltenyi). Purity was routinely checked to >80% of alive cells. Feeder cells were obtained from the 4-1BB-PE negative fraction by CD3 MicroBead (Miltenyi) depletion and subsequent irradiation at 50Gy. Enriched CD40L^+^ 4-1BB^+^ cells were co-cultured with feeder cells at a ratio of 1:1 in AB-medium supplemented with 10 ng/ml IL-7 and IL-15 respectively (both from Miltenyi) for 10 d followed by 2 d cytokine starvation. Restimulation in the presence of CD3 depleted, autologous APC was conducted as described above and indicated in the figure legends. For spike glycoprotein epitope identification restimulation was performed with the Epitope Mapping Peptide Set SARS-CoV-2 (JPT) according to the manufacturer’s instructions.

### Flow Cytometry

Stimulations were stopped by incubation in 20 mM EDTA for 5 min. Surface staining was performed for 15 min in the presence of 1 mg/ml Beriglobin (CSL Behring) with the following fluorochrome-conjugated antibodies titrated to their optimal concentrations: CD3-FITC (REA613, Miltenyi), CD4-VioGreen (REA623, Miltenyi), CD8-VioBlue (REA734, Miltenyi), CD38-APC (REA671, Miltenyi), HLA-DR-PerCpVio700 (REA805, Miltenyi). During the last 10 min of incubation, Zombie Yellow fixable viability staining (Biolegend) was added. Fixation and permeabilization were performed with eBioscience™ FoxP3 fixation and PermBuffer (Invitrogen) according to the manufacturer’s protocol. Intracellular staining was carried out for 30 min in the dark at room temperature with 4-1BB-PE (REA765, Miltenyi) and CD40L-PeVio770 (REA238, Miltenyi). All samples were measured on a MACSQuant^®^ Analyzer 16 (Miltenyi). Instrument performance was monitored prior to every measurement with Rainbow Calibration Particles (BD).

### SARS-CoV-2 IgG S1 ELISA

Anti-SARS-CoV-2 IgG and IgA ELISA specific for S subunit 1 (S1) using a commercial kit (EUROIMMUN Medizinische Labordiagnostika AG) were performed according to the manufacturer’s instructions and described previously (*40*). Maximum values were set as 10 for IgA.

### SARS-CoV-2 Neutralization Assay

Neutralization activity of SARS-CoV-2 specific antibodies was assessed with a plaque reduction neutralization test (PRNT) as described before (*39*). Vero E6 cells (1.6 ×10^5^ cells/well) were seeded in 24-well plates overnight. Sera were diluted in OptiPro and mixed 1:1 with 200µl virus (Munich isolate 984) solution containing 100 plaque forming units and incubated at 37°C for 1 hour. Plated Vero cells were incubated with 200µl serum-virus solution. After 1 hour at 37°C, the supernatants were discarded and cells were washed once with PBS and overlaid with 1.2% Avicel solution in DMEM. After 3 days at 37°C, the supernatants were removed, the cells were fixed with 6% formaldehyde in PBS and stained with crystal violet. All dilutions were tested in duplicates.

### Epitope specific antibody ELISA

400 nM of biotinylated peptide 204_-1(Biotin-Ttds-PSKPSKR*SFIEDLLFNKV*-OH, overlapping with 204_3 by 11 aa (cursive letters), Ttds linker = N-(3-{2-[2-(3-Aminopropoxy)-ethoxy]-ethoxy}-propyl)-succinamic acid, JPT Peptide Technologies) was coated on a 96 well Streptavidin plate (Steffens Biotechnische Analysen GmbH) for 1 h at RT. After blocking (1 h, 30°C) serum samples were diluted 1:100 and incubated for 1 h at 30°C. HRP-coupled, anti-human-IgG secondary antibody (Jackson Immunoresearch) was diluted 1:5000 (Jackson Immunoresearch) and added to the serum samples for 1 h at 30°C, then HRP substrate was added (TMB, Kem-En-Tec). The reaction was stopped by adding sulfuric acid and absorption was measured at 450 nm using a FlexStation 3.

### HLA typing and analysis

HLA typing has been performed by LABType® CWD assays (One Lambda, West Hills, CA, USA) based on reverse sequence-specific oligonucleotides (rSSO) according to the manufacturer’s instructions. Briefly, the HLA genomic region has been amplified individually using locus-specific biotinylated primers for HLA-DRB1, -DQA1, -DQB1, -DPA1 and –DPB1. Amplicons were hybridized to HLA allele- and allele-group-specific probes attached to Luminex® beads. Complementary binding was detected by addition of R-phycoerythrin-conjugated streptavidin and acquired using a FLEXMAP 3D flow analyzer (Luminex, Austin, TX, USA). HLA typings were derived as 2-field typing results with the highest probability as referenced in the catalogue of common and well-documented HLA alleles version 2.0.0 33. IEDB Analysis Resource (www.IEDB.org (*41, 42*), 2.22 method) was used for MHC class II binding prediction of the peptide 204_3 and homologous HCoV epitopes. For the purpose of this analysis, we refer to an individual as ‘homozygous’ if the two corresponding alleles of the same locus are identical in the first two fields.

### Homology score

For the calculation of the homology score all possible 9mers were generated for each respective PepMix of SARS-CoV-2. Each of the 9mers was scored against each unique 9mer from the proteomes of the corona viruses 229E, NL63, OC43 and HKU1 (isolates N1, N2, N5) using the PAM30 substitution matrix. The homology score is the percentage of comparisons with a pairwise 9mer-score above 30.

### Data analysis and statistics

Study data were collected and managed using REDCap electronic data capture tools hosted at Charité (*43, 44*). Flow cytometry data were analyzed with FlowJo 10.6 (FlowJo LLC) and statistical analysis conducted with GraphPad Prism 9. If not stated otherwise, data are plotted as mean. *N* indicates the number of donors. P-values were set as follows: *p<0.05, ** p<0.01, and *** p<0.001.

---

**Fig. S1:**
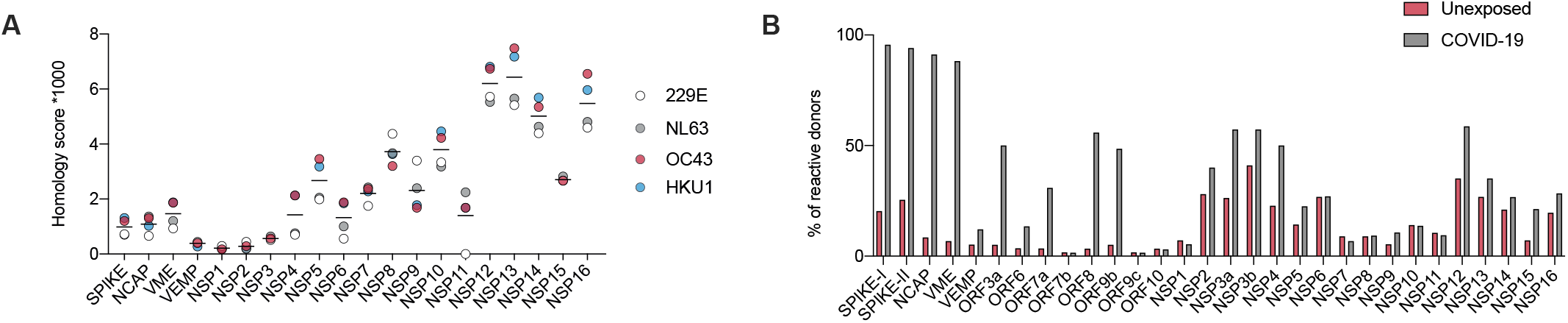
HCoV homology scores and SARS-CoV-2 peptide pool reactivity. **(A)** Similarity between SARS-CoV-2 and the different endemic coronaviruses 229E, NL63, OC43 and HKU1 (isolate N1). Scores were calculated using a PAM30 substitution matrix for all 9mers of the respective antigen against all 9mers of the proteome of SARS-CoV-2. The homology score is the percentage of 9mer pairs with a score above 30. **(B)** Frequency of donors with reactive T cells against indicated peptide pools in unexposed and COVID-19 convalescents from Fig. 1A with a stim. index ≥ 3.

**Fig. S2:**
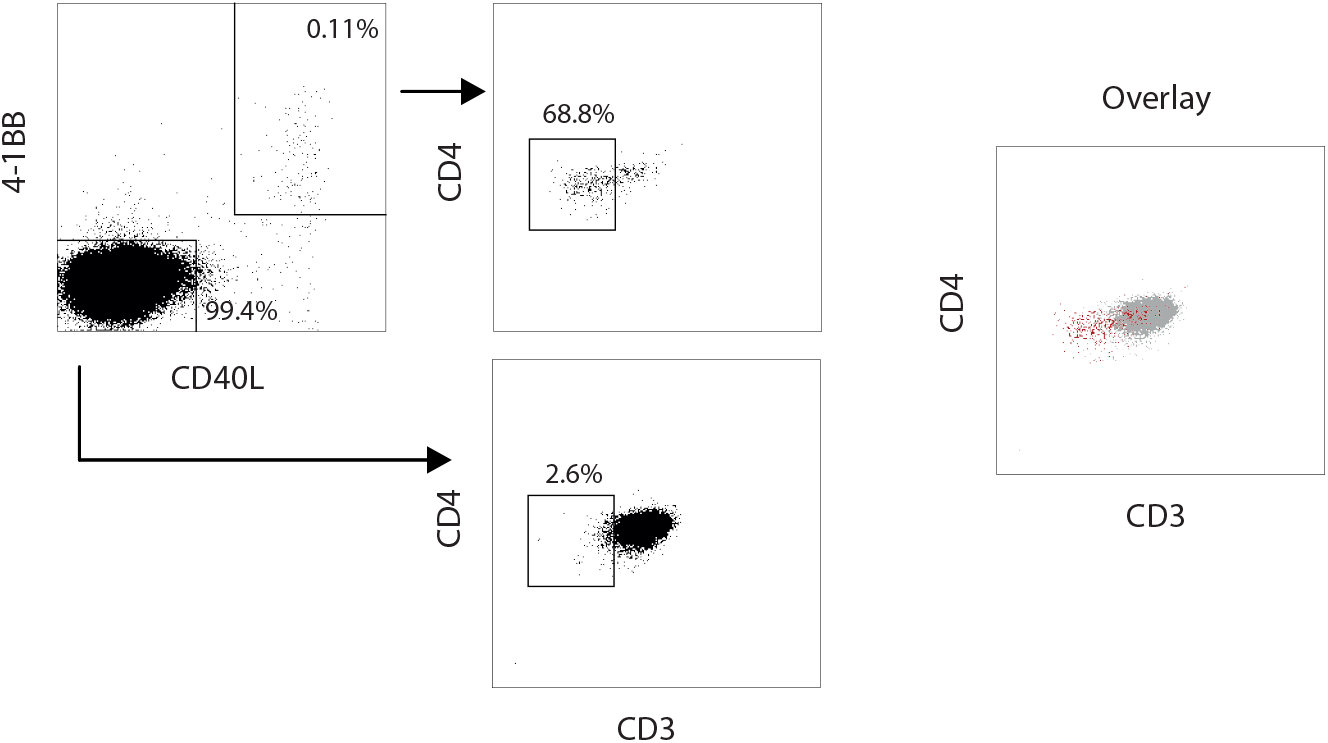
CD3^low^ gating in flow cytometry. Representative dot plot of gating for CD3^low^ cells within non-reactive CD4^+^ T cells and S-II-reactive CD40L^+^ 4-1BB^+^ CD4^+^ T cells. Cells were pre-gated on alive, doublet-free CD4^+^ lymphocytes.

**Fig. S3:**
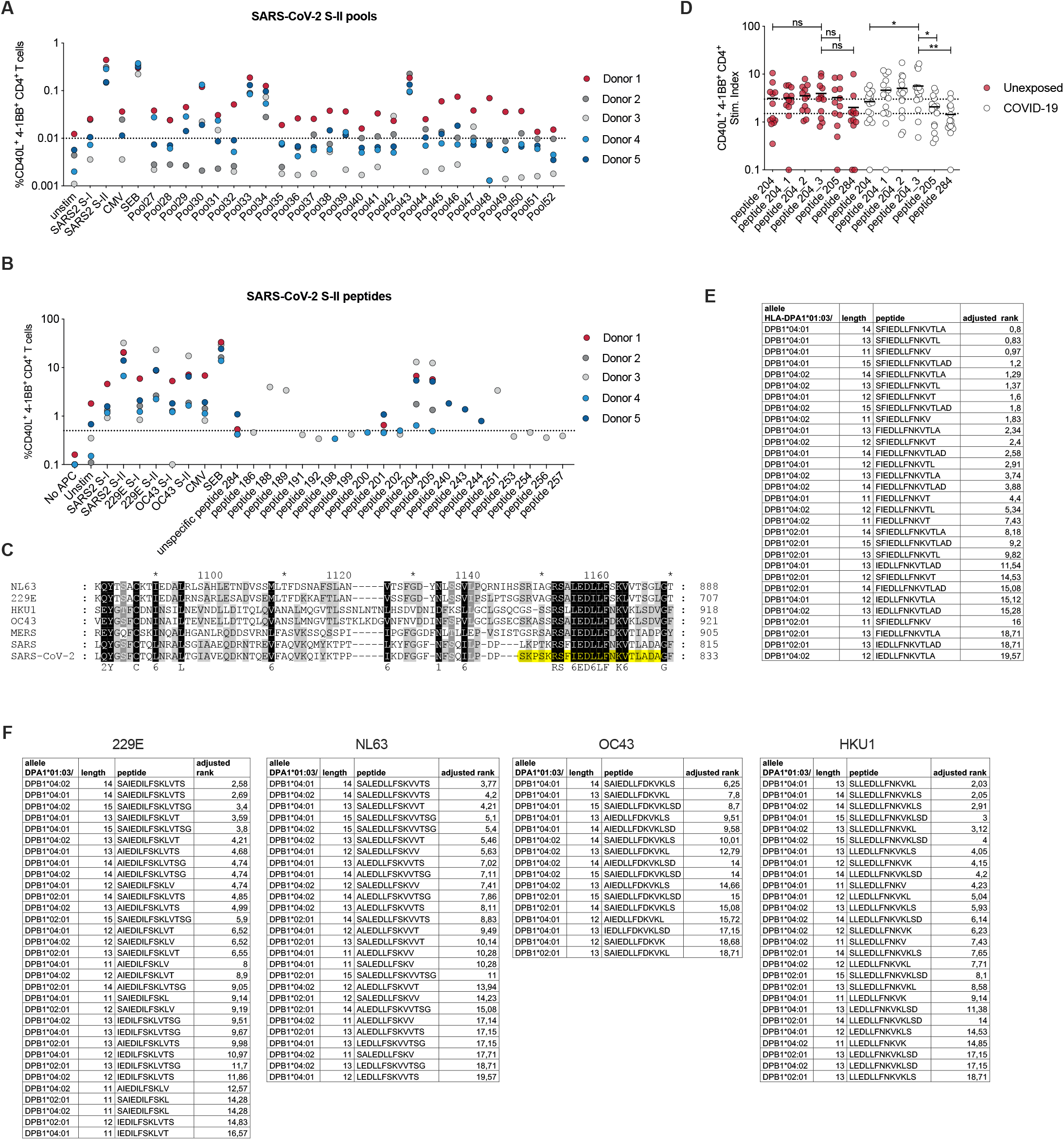
Identification of *iCope* by narrowing down the sequence regions mapped in SARS-CoV-2 peptide pools that exhibited highest T cell cross-reactivity. **(A)** Expanded SARS-CoV-2 S-I or S-II specific CD40L^+^ 4-1BB^+^ CD4^+^ short-term-culture T cells were restimulated with autologous APC in the presence of different matrix pools and the frequency of CD40L^+^ 4-1BB^+^ reactive cells determined. Cut-off line is set at 0.01 based on unstimulated controls. **(B)** Expanded SARS-CoV-2 S-I or S-II reactive CD40L^+^ 4-1BB^+^ CD4^+^ T cells were restimulated with autologous APC in the presence of single peptide candidates determined from matrix stimulation (shown in A) and the frequency CD40L^+^ 4-1BB^+^ reactive CD4^+^ T cells was determined. Cut-off line is set at 0.5 based on unstimulated controls. **(C)** Position of single peptides 204 and 205 (yellow) in sequence overlay of the known 7 human coronaviruses. Color code indicates homology: black = identical amino acid, grey = conservative amino acid replacement. **(D)** Identification of the dominant peptide at the 204 to 205 sequence intersection. The number behind the underline character indicates the number of amino acids by which the 15-mer was shifted towards the C-terminus beginning with the sequence of 204 and ending with the sequence of 205. Only a selection of donors is shown that displayed a stim. index ≥ 3 for a minimum of one stimulation. **(E, F)** MHC-II HLA binding prediction of HLA-DPA1*01:03/DPB1*02:01, HLA-DPA1*01:03/DPB1*04:01 and HLA-DPA1*01:03/DPB1*04:02 with *iCope* and fragments thereof (E) or HCoV derived homologous peptides and fragments thereof (F).

**Fig. S4:**
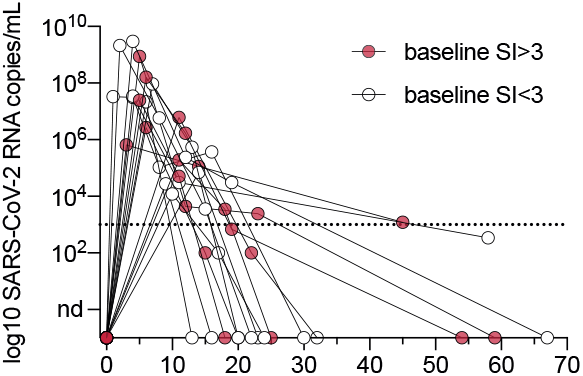
SARS-CoV-2 RNA concentration kinetics. Viral RNA copies per mL diluted swab at indicated time points. Dotted line indicates the limit of quantification. nd, no SARS-CoV-2 RNA detection in this sample. Donors with stim. Index ≥ 3 for SARS-CoV-2 S-II at baseline shown as red circles, individuals with stim. index ≤ 3 shown with white circles.

**Fig. S5:**
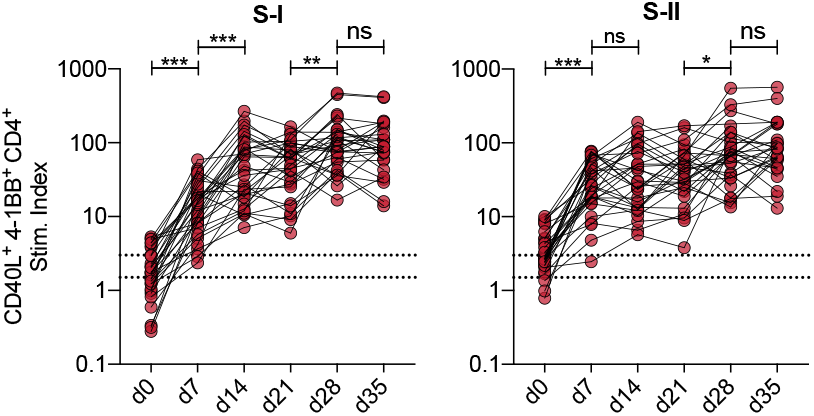
Individual T cell responses to vaccination. Individual vaccinee response kinetics of CD40L^+^ 4-1BB^+^ CD4^+^ T cells upon stimulation with S-I or S-II, as shown in Fig. 6B. *p<0.05, **p<0.01, ***p<0.001 and ns for p>0.05 (Paired Student’s *t*-test). Secondary vaccination occurred on day 21.

